# Longitudinal Changes in COVID-19 Associated In-Hospital Mortality

**DOI:** 10.1101/2021.05.04.21255938

**Authors:** Frederick Warner, Matt D.T. Hitchings, Derek A.T. Cummings, Jacob McPadden, Harlan M. Krumholz, Albert I. Ko, Wade L. Schulz

## Abstract

**Objective:** As the COVID-19 pandemic has evolved, a key question for health care systems is whether in-hospital mortality has changed over time and if so, what factors contributed to these changes. Our goal was to leverage real-world data spanning two COVID-19 surges over the first year of the pandemic to determine the temporal trend of in-hospital mortality.

**Design:** This was an observational, retrospective study based on real-world data for patients admitted with COVID-19. Generalized additive models (GAM) were used to evaluate the association of covariates with the composite outcome over time.

**Setting and Population:** We identified a retrospective cohort of all patients who were hospitalized within the Yale New Haven Health (YNHH) system with an admission diagnosis of COVID-19 between March 1, 2020 and February 28, 2021.

**Main outcome:** The primary outcome for the study was a composite endpoint of in-hospital mortality, defined as death during the index hospitalization or discharge to hospice.

**Results:** Among 6,477 discharges over the study period, the mean age was 66.2 years (SD=17.6), 52.5% (n=3,401) were male and the overall composite mortality was 14.2% (n=920). Composite in-hospital mortality was significantly associated with increased age, comorbidity index, respiratory rate, and heart rate; decreased systolic blood pressure; male sex; and admission from a long-term care facility (LTCF). The significant temporal decrease in mortality that was observed for patients admitted from a location other than a LTCF was not seen in those admitted from a LTCF.

**Conclusions:** We found that the adjusted in-hospital mortality rate declined over the first year of the pandemic, despite a second surge in COVID-19-related hospitalizations. Importantly, the decrease in mortality appeared to be driven by declines in risk in those not admitted from a LTCF. The observed decrease in mortality over time suggests that improved outcomes may be due to progressive, incremental learning and continuous evolution in hospital practice and policy over the course of the pandemic.

## Background

As the COVID-19 pandemic has evolved, a key question for health care systems is whether in-hospital mortality has changed over time and if so, what factors contributed to these changes. Reports from several settings have observed decreases in mortality [1–3] and suggested that these decreases were not attributable to temporal changes in patient characteristics [2]. However, these studies reported data over a limited time period that encompassed a single surge of COVID-19 cases.

Our goal was to leverage real-world data spanning two COVID-19 surges within a one-year period to determine the temporal trend of in-hospital mortality. We used electronic health record data to obtain detailed clinical information and evaluate a range of time-varying explanatory factors for observed trends, which included patient comorbidity burden, admission from long-term care facilities (LTCFs), and disease severity at the time of hospital admission.

## Methods

We identified a retrospective cohort of patients who were hospitalized within the Yale New Haven Health (YNHH) system with an admission diagnosis of COVID-19 as previously described [4] (Supplemental Methods) between March 1, 2020 and February 28, 2021. Data were extracted on March 26, 2021 from our local Observational Medical Outcomes Partnership (OMOP) data repository and analyzed within our computational health platform [5]. Patients who remained admitted to the hospital at the time of the data extract were excluded from analysis. For patients with multiple COVID-19 admissions, only the index (first) admission was considered for analysis.

The primary outcome for the study was a composite endpoint of in-hospital mortality, defined as death during the index hospitalization or discharge to hospice. Generalized additive models (GAM) were used to evaluate the association of covariates with the composite outcome over time. Covariates included demographic data (age, gender, race, ethnicity), first-recorded vital signs (respiratory rate, systolic blood pressure, heart rate), comorbidities prior to admission, admission from a LTCF, and day of hospital admission after March 1, 2020. Comorbidity burden was calculated from an Elixhauser comorbidity analysis as a weighted score using the van Walraven algorithm [6].

## Results

Among 6,493 COVID-19 patients who were admitted to YNHH between March 1, 2020 and February 28, 2021, 6,477 (99.8%) were discharged at the time of analysis (Figure 1A). Of discharged patients, the mean age was 66.2 years (SD=17.6), 52.5% (n=3,401) were male, 56.3% (n = 3,647) had a recorded race of White, 21.2% (n=1,374) had a recorded race of Black, 22.3% (n=1,446) had a recorded ethnicity of Hispanic or Latino, and 10.1% (n=656) were admitted from a LTCF. Overall composite mortality was 14.2% (n=920).

**Figure 1.**
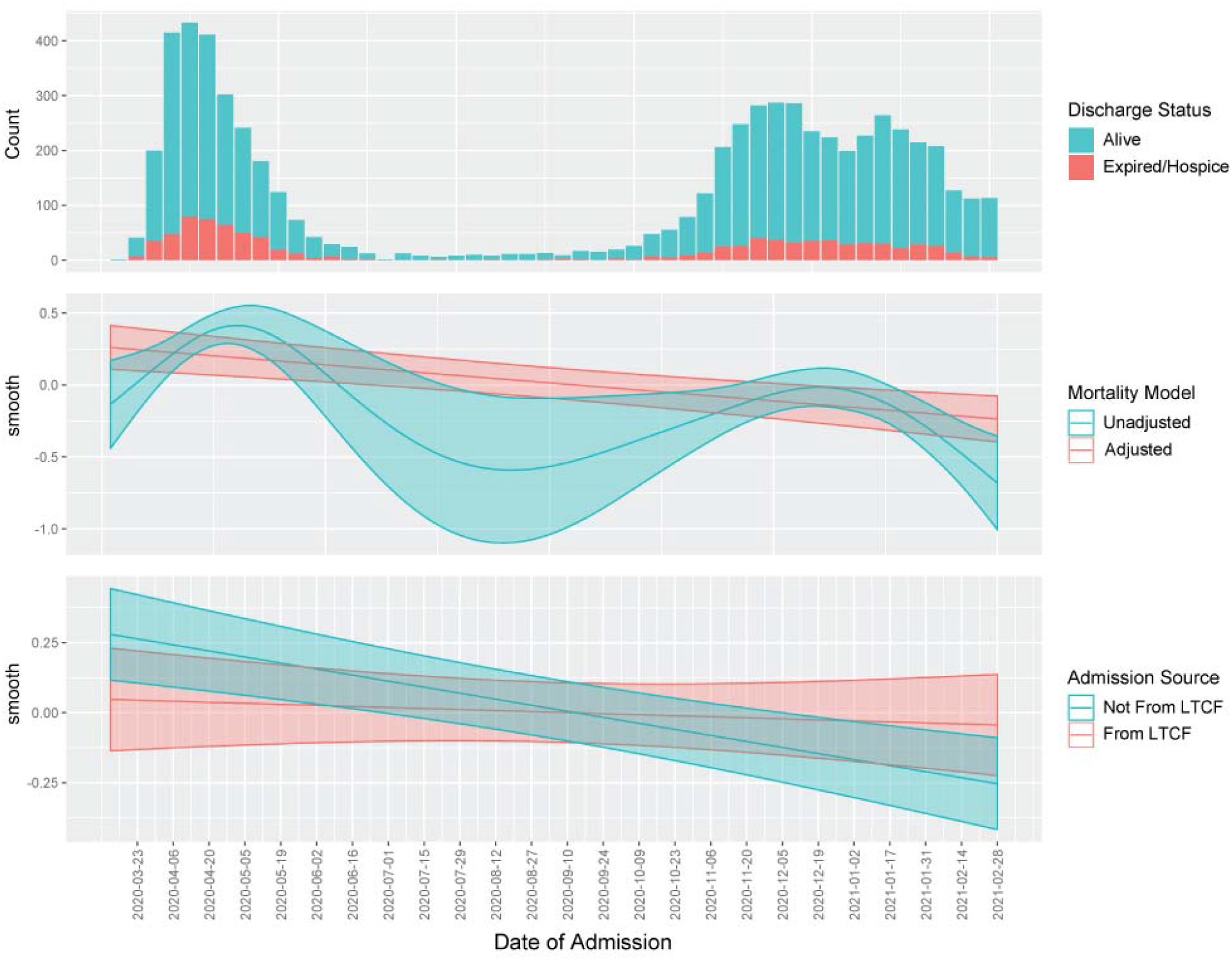
In-hospital outcomes for COVID-19 patients admitted between March 1, 2020 and February 28, 2021. A) Number of discharged patients based on date of admission who had the composite endpoint of death or hospice (orange) or were discharged alive (blue). B) Unadjusted and adjusted smooth terms (+2 standard errors) for the interaction between day of admission and the composite endpoint during the time series. C) Adjusted smooth terms (+2 standard errors) stratified according to admission from a long-term care facility (LTCF, orange line) or other source (blue lines).

Composite in-hospital mortality was significantly associated with increased age, comorbidity index, respiratory rate, and heart rate; decreased systolic blood pressure; male sex; and admission from a LTCF (Table 1). After adjusting for other covariates, the mortality smooth term for day declined linearly (Figure 1B). The significant temporal decrease in mortality that was observed for patients who were not admitted from a LTCF was not seen in those admitted from a LTCF (Figure 1C). The only other covariate for which we identified a significant interaction with day was a recorded race of White.

**Table 1.** Generalized additive models of in-hospital mortality. Generalized additive models were used to estimate coefficients and estimated degrees of freedom (EDFs) of categorical and continuous covariates, respectively, for unadjusted mortality over time (Model 1), adjusted mortality over time (Model 2), and adjusted mortality over time which includes an interaction term for day and admission source (Model 3). The adjusted models omit 21 of 6,477 individuals due to missing vital sign data.

|  | Unadjusted Mortality |  | Adjusted Mortality |  |  |  |
| --- | --- | --- | --- | --- | --- | --- |
|  | *Model 1* |  | *Model 2* |  | *Model 3* |  |
|  | *Est* | *Pr(>\|z\|)* | *Est* | *Pr(>\|z\|)* | *Est* | *Pr(>\|z\|)* |
| **Coefficients** |  |  |  |  |  |  |
| (Intercept) | -1.82 | <2e-16 | -2.43 | <2e-16 | -2.43 | <2e-16 |
| Male |  |  | 0.24 | 0.003 | 0.24 | 0.003 |
| White |  |  | -0.22 | 0.195 | -0.22 | 0.194 |
| Asian |  |  | 0.57 | 0.070 | 0.59 | 0.063 |
| Black or African American |  |  | -0.15 | 0.441 | -0.15 | 0.448 |
| Hispanic |  |  | 0.10 | 0.565 | 0.10 | 0.545 |
| Admission from LTCF |  |  | 0.63 | <0.001 | 0.71 | <0.001 |
| **Smooth Terms** | *EDF* | *p* | *EDF* | *p* | *EDF* | *p* |
| Day | 5.10 | <0.001 | 0.95 | <0.001 |  |  |
| Interaction with Non-LTCF admission |  |  |  |  | 0.95 | <0.001 |
| Interaction with LTCF admission |  |  |  |  | 0.28 | 0.242 |
| Age |  |  | 1.00 | <2e-16 | 1.00 | <2e-16 |
| Comorbidity Score |  |  | 0.98 | <0.001 | 0.98 | <0.001 |
| Initial Heart Rate |  |  | 3.33 | 0.002 | 3.19 | 0.002 |
| Initial Systolic Blood Pressure |  |  | 2.10 | <0.001 | 2.11 | <0.001 |
| Initial Respiratory Rate |  |  | 3.71 | <2e-16 | 3.67 | <2e-16 |

## Discussion

We found that the in-hospital mortality rate declined over the first year of the pandemic, despite a second surge in COVID-19 related hospitalizations. After adjusting for covariation in patient demographics and disease severity on admission over time, we found the decrease in mortality rate to be progressive without an increase in adjusted mortality rate during the second peak of the pandemic. Importantly, the decrease in mortality appeared to be driven by declines in risk for patients not admitted from a LTCF, suggesting that patients from LTCFs may have additional and ongoing risks for COVID-19 related mortality.

This study has several limitations including the observational study design and potential for unmeasured confounding. Secondly, data related to treatments were not included in the analysis. We therefore could not ascertain whether temporal changes in mortality were due to changes in treatment practices. Thirdly, this study represents findings from a single health system which may limit the generalizability of our findings. Finally, we did not evaluate COVID-19 associated mortality after discharge from the index hospitalization.

Overall, the observed decrease in mortality over time suggests that improved outcomes may be due to progressive, incremental learning and continuous evolution in hospital practice and policy over the course of the pandemic. While we adjusted for some risks, including overall comorbidity burden, it is possible that specific underlying conditions in those admitted from long-term care may be associated with worse outcomes. Additional studies will be needed to identify the causative factors for the decline in mortality and the reasons why mortality rates did not decline in those admitted from LTCFs.

## Data Availability

Data cannot be shared publicly because of the presence of potentially identifiable health information. Requests for access can be directed to the corresponding author and Yale Human Research Protection Program.

## Disclosures

A.I.K. reports grants from Bristol Myer Squib, grants from Regeneron, grants from Serimmune, grants and personal fees from Tata Medical Devices outside the submitted work. H.M.K. works under contract with the Centers for Medicare & Medicaid Services to support quality measurement programs, was a recipient of a research grant from Johnson & Johnson, through Yale University, to support clinical trial data sharing; was a recipient of a research agreement, through Yale University, from the Shenzhen Center for Health Information for work to advance intelligent disease prevention and health promotion; collaborates with the National Center for Cardiovascular Diseases in Beijing; receives payment from the Arnold & Porter Law Firm for work related to the Sanofi clopidogrel litigation, from the Martin Baughman Law Firm for work related to the Cook Celect IVC filter litigation, and from the Siegfried and Jensen Law Firm for work related to Vioxx litigation; chairs a Cardiac Scientific Advisory Board for UnitedHealth; was a member of the IBM Watson Health Life Sciences Board; is a member of the Advisory Board for Element Science, the Advisory Board for Facebook, and the Physician Advisory Board for Aetna; and is the co-founder of Hugo Health, a personal health information platform, and cofounder of Refactor Health, a healthcare AI-augmented data management company. W.L.S. was an investigator for a research agreement, through Yale University, from the Shenzhen Center for Health Information for work to advance intelligent disease prevention and health promotion; collaborates with the National Center for Cardiovascular Diseases in Beijing; is a technical consultant to Hugo Health, a personal health information platform; cofounder of Refactor Health, an AI-augmented data management platform for healthcare; and was a consultant for Interpace Diagnostics Group, a molecular diagnostics company. The other authors have no relevant disclosures.

## Notes

### Funding Statement

No external funding was received for this work.

### Author Declarations

The study was approved by the Yale University Institutional Review Board (IRB #2000027975).

### Summary of Updates

Reference added for cohort selection criteria

