## Supplemental Methods for "Longitudinal Changes in COVID-19 Associated In-Hospital Mortality"

*Data Sources:* This was an observational, retrospective study of patients who were admitted to the Yale New Haven Health (YNHH) system with a diagnosis of SARS-CoV-2 infection. YNHH uses a single electronic health record (EHR) across a health system with five inpatient sites, including pediatric, suburban community, urban community, and urban academic centers. Demographics, comorbidities, vital signs and visit outcomes were extracted on March 26, 2021 from our local Observational Medical Outcomes Partnership (OMOP) data repository and analyzed within our computational health platform. The study was approved by the Yale University Institutional Review Board (IRB #2000027975).

*Cohort Definition:* The study cohort consists of all patients in the YNHH system who were admitted with a SARS-CoV-2 diagnosis before March 1, 2021. A diagnosis of SARS-CoV-2 was made using two distinct criteria, both using ICD-10-CM coding. The first criterion was that an admitted patient had a principal diagnosis of either U07.1, J12.81, or B97.21. Additionally, the criterion accepted B97.29 as a principal diagnosis if coded prior to April 1, 2020. The second criterion was designed to capture patients with severe Covid-19 symptoms, the source of which was considered to be underlying SARS-CoV-2 infection. Under this criterion, admitted patients were required to have a secondary admit diagnosis of U07.1, along with a primary diagnosis of any of the following codes: A41.89, A41.9, J12.89, J96.01, J18.9, J80, B34.2, J96.21 or J06.9. For patients with multiple SARS-CoV-2 admissions during the study period, only the index (first) admission was considered for analysis.

*Study Outcome:* The outcome for the study was a composite discharge disposition combining patients who expired in-hospital with those sent to hospice (both facility-based and in-home), an outcome which is consistent with that employed in related studies. For patients with two or more SARS-CoV-2 admissions, the discharge disposition of an earlier visit was inherited from a later visit if the start time of the later visit was within 3 hours of the end time of the earlier visit. Patients who were still admitted to the hospital at the time of the data extract were excluded from analysis.

*Statistical Analyses:* A Generalized Additive Model (GAM) analysis was performed to identify risk factors associated with having the study (composite) outcome of expired or hospice discharge status. Risk factors studied included demographic data (age, gender, race, ethnicity), first-recorded vital signs (respiratory rate, blood pressure, heart rate), the day of admission, overall comorbidity level, and admission status. The day of admission was given in days after March 1, 2020, and the comorbidity level was calculated from an Elixhauser comorbidity analysis as a weighted score using the van Walraven algorithm. Admission status ("admit status") refers to whether the patient was admitted from a skilled nursing facility, intermediate care facility, assisted living facility, or hospice versus all other admission sources.

All statistical work was done within our computational health platform using R (version 3.5.1). Data were extracted from the OMOP data repository using the sparklyr package (version 1.2.0), and the Elixhauser comorbidity data was computed using the comorbidity package (version 0.5.3). The mgcv package (version 1.8-29) was used for the GAM analysis.
